# Diagnosing Appendicitis with Precision: A Comparative Analysis of Modified Alvarado and RIPASA Scoring Systems in a Northern Divisional Hospital of Bangladesh

**DOI:** 10.1101/2024.11.06.24316874

**Authors:** Nafisa Naz, Md Mostafa Monower, Shah Md Ahsan Shahid, Syeda Momena Hossain, Tanvir Ahmad, Baharul Islam

## Abstract

**Objective:** This study aimed to evaluate the diagnostic efficacy of the Modified Alvarado Scoring System (MASS) and the Raja Isteri Pengiran Anak Saleha Appendicitis (RIPASA) scoring system, assessing their sensitivity, specificity, positive predictive value (PPV), negative predictive value (NPV), and overall accuracy.

**Methods:** A cross-sectional analysis was conducted at the Department of General Surgery, Rajshahi Medical College Hospital, Bangladesh, from September to December 2020. The study included 138 purposively selected individuals aged 13 years and above, suspected of acute appendicitis. Data were collected through structured interviews, detailing socio-demographic characteristics, medical history, clinical examinations, and specific laboratory tests (CBC, Urine RE). Histopathology reports from post-operative cases were used as the gold standard for diagnosis. MASS and RIPASA scores, derived from their respective criteria, were analyzed using STATA.

**Results:** Participants had a mean age of 26.2 years, with males constituting 55.0% of the sample. The MASS scoring system reported a sensitivity of 79.8%, specificity of 57.9%, PPV of 92.2%, NPV of 31.4%, and an overall diagnostic accuracy of 76.8%. In contrast, the RIPASA scoring system demonstrated a sensitivity of 96.6%, specificity of 73.7%, PPV of 95.8%, NPV of 77.8%, and a diagnostic accuracy of 93.5%. ROC AUC analysis yielded values of 0.6886 for MASS and 0.8516 for RIPASA, indicating a statistically significant difference (p<0.05).

**Conclusion:** The findings highlight the superior clinical utility of the RIPASA scoring system over MASS, particularly in settings with limited access to advanced diagnostic facilities. Adopting the RIPASA scoring system could significantly enhance the diagnosis of acute appendicitis, suggesting its potential for improving clinical outcomes in similar healthcare environments.

## Introduction

Acute appendicitis is a prevalent surgical emergency that demands prompt and accurate diagnosis for optimal patient management.[1] However, the diagnostic challenge of acute appendicitis is compounded by atypical symptoms in approximately 50% of cases and the potential for other conditions to mimic its signs and symptoms.[2] Medical history, clinical examination, and laboratory investigations are critical to the diagnostic process. Despite the availability of diagnostic aids such as abdominal Ultrasonography, Computerized Tomography, Laparoscopy, Magnetic Resonance Imaging, and Computer-Aided Barium Enema, their utility is limited by the need for expertise and associated high costs.[3] In peripheral districts, limited access to these diagnostic facilities further complicates the accurate diagnosis of acute appendicitis, underscoring the need for accurate and timely diagnosis to facilitate appropriate patient management and optimize clinical outcome.[4]

To aid in the diagnosis of acute appendicitis, various scoring systems have been developed to improve diagnostic accuracy and reduce unnecessary surgical interventions.[5] The Alvarado scoring system, introduced in 1986 and later refined as the Modified Alvarado Scoring System (MASS) in 1994,[6], [7] assigns scores based on clinical signs, symptoms, and laboratory findings associated with acute appendicitis. It has been extensively studied and validated across different populations, demonstrating reasonable accuracy in diagnosing appendicitis. Conversely, the Raja Isteri Pengiran Anak Saleha Appendicitis (RIPASA) scoring system, developed by Chong et al. in 2010,[2] is tailored specifically for the Asian population. It includes clinical, laboratory, and imaging findings alongside demographic characteristics, offering promising results in diagnostic accuracy and potential advantages over other scoring systems.[8], [9]

Despite the widespread application of both scoring systems, a consensus on their comparative performance in various clinical settings, especially in resource-limited environments like peripheral district hospitals, remains elusive.[10] Moreover, research directly comparing the Modified Alvarado (MASS) and RIPASA Scoring Systems within the context of Bangladesh’s peripheral medical facilities is scarce. Therefore, this study aims to conduct a thorough analysis of both scoring systems’ diagnostic performance, assessing their sensitivity, specificity, positive predictive value (PPV), negative predictive value (NPV), and overall accuracy. The insights gained from this research are expected to contribute to minimizing negative appendectomies, particularly in peripheral hospital settings.[11] The outcomes of this study may have important implications for clinical practice, guiding the selection and implementation of the most suitable scoring system for the accurate and efficient diagnosis of acute appendicitis in similar healthcare environments.

## Methodology

### Study design, setting and sampling

This cross-sectional study was conducted at the Department of General Surgery, Rajshahi Medical College Hospital, Bangladesh, from September to December 2020. A total of 173 patients suspected of acute appendicitis were initially considered for the study (**Supplementary figure 1**). However, 35 patients (20.2%) were excluded due to unavailable histopathology reports (n=17), conservative treatment (n=12), and urogenital causes identified through ultrasonography (n=6). After these exclusions, the final sample size was 138 patients. Additional exclusions were applied to patients with a prior history of RIF pain, those who developed RIF pain after being admitted for another reason, or those with generalized peritonitis, septic shock, or pregnancy.

### Data collection

Data on suspected acute appendicitis patients were collected through investigator-led interviews in the admission unit.[12] Informed consent was obtained prior to 15 to 20-minute sessions, which covered demographic, and medical history details. Clinical examination data were recorded at admission or retrieved from post-operative files. Routine laboratory investigations (CBC, Urine RME) and histopathology reports of post-operative cases were collected for further analysis. Surgical decisions were made by the attending surgeons. A ward round the following morning facilitated the completion of pending information and enrollment of new cases. Data collection utilized a pre-tested, semi-structured questionnaire available in both English and Bangla.

### *Index* case

In this study, the Modified Alvarado Scoring System (MASS) and the Raja Isteri Pengiran Anak Saleha Appendicitis (RIPASA) scoring system were used as the index tests to predict acute appendicitis. The MASS score was calculated based on seven standard clinical and laboratory parameters, while the RIPASA score utilized 14 parameters, including an additional criterion for patients holding a Foreign National Record of Identity Card (NRIC), as outlined in **Supplementary Tables 1 and 2**, respectively. To minimize bias in surgical decision-making, neither scoring system was calculated during the data collection phase. Instead, the total scores were categorized post-hoc using a cut-off threshold of 7 for MASS and 7.5 for RIPASA, with scores above these thresholds indicating a high likelihood of appendicitis.

### Gold standard

In this study, histopathology served as the operational gold standard for confirming acute appendicitis of the post-operative cases. It ensured a definitive and accurate assessment of the condition, providing a detailed understanding of the underlying pathological changes. Consistent with findings from the literature, where histopathology has been widely acknowledged as the gold standard for confirming acute appendicitis,[9], [13], [14], [15], [16] post-operative specimens’ histopathological analysis was considered the benchmark for confirming cases of acute appendicitis and distinguishing them from normal or non-appendicitis cases.

### Quality control

The study implemented extensive quality control measures, including pre-piloting of the questionnaire and daily review of completed forms for consistency and completeness. Laboratory data and histopathological reports were verified against patient identifiers to ensure accuracy. The research adhered strictly to its planned timeline and budget.

### Minimization of bias

Several steps were taken to minimize potential sources of bias. To reduce selection bias, participants were purposively selected, though this may limit the generalizability of the findings. Recall bias was addressed by ensuring that data were collected through structured interviews conducted shortly after admission, reducing the reliance on long-term memory. Observer bias was minimized by using consistent clinical examination protocols across all surgeons. Additionally, neither the Modified Alvarado nor the RIPASA scores were calculated during data collection to prevent decision-making bias in surgical procedures.

### Statistical analysis

Descriptive statistics were used to summarize the socio-demographic and clinical characteristics of the participants. The diagnostic performance of the Modified Alvarado Scoring System (MASS) and the Raja Isteri Pengiran Anak Saleha Appendicitis (RIPASA) scoring system was assessed by calculating their sensitivity, specificity, positive predictive value (PPV), negative predictive value (NPV), and overall diagnostic accuracy, using histopathological findings as the reference standard.

Receiver Operating Characteristic (ROC) curves were generated for both scoring systems by systematically varying the classification thresholds. The Area Under the ROC Curve (AUC) was determined to evaluate each system’s discriminative capability in diagnosing appendicitis. Statistical associations between categorical variables were assessed using the chi-square (χ2) test. All p-values were two-tailed, with statistical significance set at p<0.05. Statistical analyses were performed using STATA 17, with findings presented in tables and graphs.

### Ethical consideration

Ethical approval for this study was granted by the Institutional Review Board (IRB) of Rajshahi Medical College Hospital (Ref: RMC/IRB/2019/20-011/54, Date: 04 Aug 2020). Detailed information about the study’s objectives, procedures, potential risks, and benefits was provided to participants before obtaining informed consent. For illiterate participants, thumbprints were collected in lieu of signatures, witnessed and co-signed. Participants were assured of their right to withdraw from the study at any time without affecting their medical care. The research was conducted in strict adherence to the Helsinki Declaration, ensuring the confidentiality and privacy of all participants. Results from medical examinations were distributed and explained to participants, who were then given the opportunity to discuss their reports with the researcher and receive appropriate guidance as needed.

## Results

The demographic characteristics, symptoms, signs, and pathological findings of the 138 participants are presented in **Table 1**. The median age was 22 years (IQR: 17, 35), with 55.0% being male. A majority (60.1%) resided in rural areas. Among symptoms, anorexia and nausea or vomiting were prevalent in 76.1% and 90.6% of cases, respectively. Notably, all participants exhibited right iliac fossa (RIF) pain, with 86.2% reporting pain extending from the umbilicus to the RIF. The duration of pain was less than 48 hours in 67.4% of cases. Clinical signs showed 47.9% had a febrile temperature, and RIF tenderness was present in all cases. Rovsing’s sign was positive in 58.0%, while muscle guarding and rebound tenderness were observed in 79.0% and 86.2% of participants, respectively. Pathological findings revealed positive urine analysis in 35.5% and leukocytosis in 88.4% of cases.

**Table 1:**
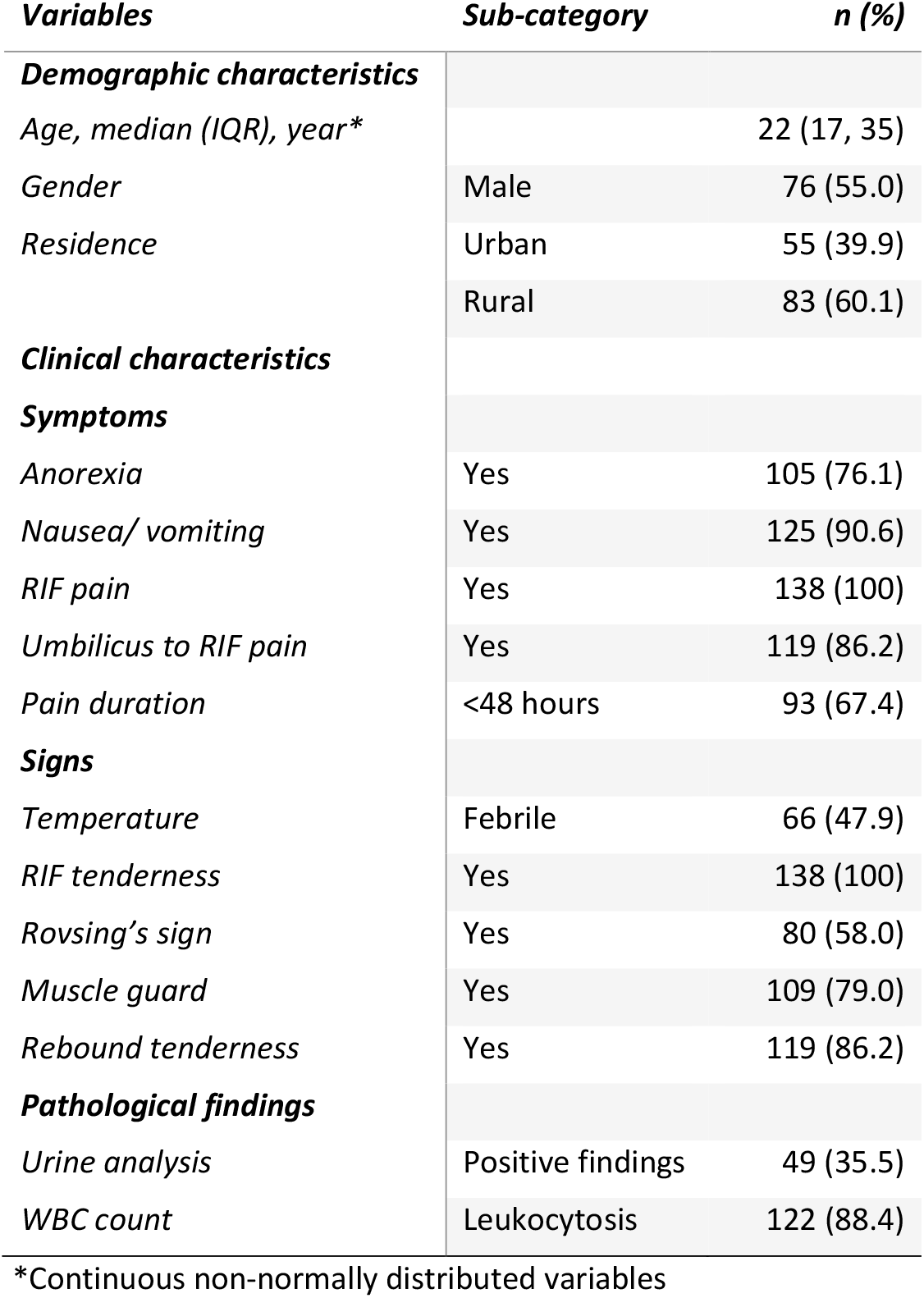
Characteristics of the participants (n=138)

| *Variables* | *Sub-category* | *n (%)* |
| --- | --- | --- |
| ***Demographic characteristics*** |  |  |
| *Age, median (IQR), year\** |  | 22 (17, 35) |
| *Gender* | Male | 76 (55.0) |
| *Residence* | Urban | 55 (39.9) |
|  | Rural | 83 (60.1) |
| ***Clinical characteristics*** |  |  |
| ***Symptoms*** |  |  |
| *Anorexia* | Yes | 105 (76.1) |
| *Nausea/ vomiting* | Yes | 125 (90.6) |
| *RIF pain* | Yes | 138 (100) |
| *Umbilicus to RIF pain* | Yes | 119 (86.2) |
| *Pain duration* | <48 hours | 93 (67.4) |
| ***Signs*** |  |  |
| *Temperature* | Febrile | 66 (47.9) |
| *RIF tenderness* | Yes | 138 (100) |
| *Rovsing's sign* | Yes | 80 (58.0) |
| *Muscle guard* | Yes | 109 (79.0) |
| *Rebound tenderness* | Yes | 119 (86.2) |
| ***Pathological findings*** |  |  |
| *Urine analysis* | Positive findings | 49 (35.5) |
| *WBC count* | Leukocytosis | 122 (88.4) |

\*Continuous non-normally distributed variables

**Table 2** details the distribution of diagnostic tool scores in relation to histopathological diagnosis. For the MASS, scores ≥7 indicated appendicitis, with 95 out of 103 cases confirmed histopathologically, and 8 classified as other conditions. For scores <7, 24 out of 35 were confirmed as appendicitis. The RIPASA utilized a cutoff ≥7.5, with 115 out of 120 cases confirmed for appendicitis, and 5 as other conditions. Scores <7.5 resulted in 4 out of 18 cases confirmed as appendicitis. Both scoring models showed significant associations with histopathological outcomes (p<0.001).

**Table 2:** Diagnostic tool scores by histopathological outcome (gold standard)

| Diagnostic tool | score | Histopathological diagnosis |  | Total | P value |
| --- | --- | --- | --- | --- | --- |
|  |  | Acute Appendicitis | Other than appendicitis |  |  |
| MASS | ≥7 | 95 | 8 | 103 |  |
|  | <7 | 24 | 11 | 35 |  |
|  | **Total** | **119** | **19** | **138** | **p<0.001** |
| RIPASA | ≥7.5 | 115 | 5 | 120 |  |
|  | <7.5 | 4 | 14 | 18 |  |
|  | **Total** | **119** | **19** | **138** | **p<0.001** |

Furthermore, **table 3** presents a comprehensive comparison of the diagnostic performance between the MASS and the RIPASA scoring system. This study identified noteworthy differences in sensitivity, specificity, positive predictive value (PPV), negative predictive value (NPV), and overall diagnostic accuracy between the two scoring systems. The MASS system demonstrated a sensitivity of 79.8% (95% CI: 71.5-86.6) and specificity of 57.9% (95% CI: 33.5-79.7), with a PPV of 92.2% (95% CI: 85.3-96.6) and an NPV of 31.4% (95% CI: 16.9-49.3), leading to a diagnostic accuracy of 76.8% (95% CI: 68.9-83.6). In contrast, the RIPASA score outperformed MASS with a sensitivity of 96.6% (95% CI: 91.6-99.1), specificity of 73.7% (95% CI: 48.8-90.9), a PPV of 95.8% (95% CI: 90.5-98.6), and an NPV of 77.8% (95% CI: 52.4-93.6), culminating in a diagnostic accuracy of 93.5% (95% CI: 88.0-97.0).

**Table 3:** Comparison of diagnostic performance between MASS and RIPASA tool.

| Parameter | MASS | RIPASA |
| --- | --- | --- |
|  | Estimate (95% C) | Estimate (95% C) |
| Sensitivity | 79.8 (71.5-86.6) | 96.6 (91.6-99.1) |
| Specificity | 57.9 (33.5-79.7) | 73.7 (48.8-90.9) |
| Positive predictive value | 92.2 (85.3-96.6) | 95.8 (90.5-98.6) |
| Negative predictive value | 31.4 (16.9-49.3) | 77.8 (52.4-93.6) |
| Diagnostic accuracy | 76.8 (68.9-83.6) | 93.5 (88.0-97.0) |

In the presented ROC AUC analysis **(Fig 1)**, this study further assessed the diagnostic performance of the MASS and RIPASA scoring systems for acute appendicitis. The ROC (Receiver Operating Characteristic) AUC (Area Under the Curve) serves as a critical metric for evaluating the discriminatory capabilities of these systems. Notably, the MASS scoring system exhibited a ROC AUC of 0.6886 (95% CI: 0.5690-0.8083), reflecting its fair capacity to distinguish between patients with and without acute appendicitis. In contrast, the RIPASA scoring system demonstrated a significantly higher ROC AUC of 0.8516 (95% CI: 0.7486-0.9546), evidencing superior discrimination between acute appendicitis and other conditions. The significant difference in performance (p<0.05) highlights RIPASA’s enhanced diagnostic accuracy in the studied population.

**Fig 1:**
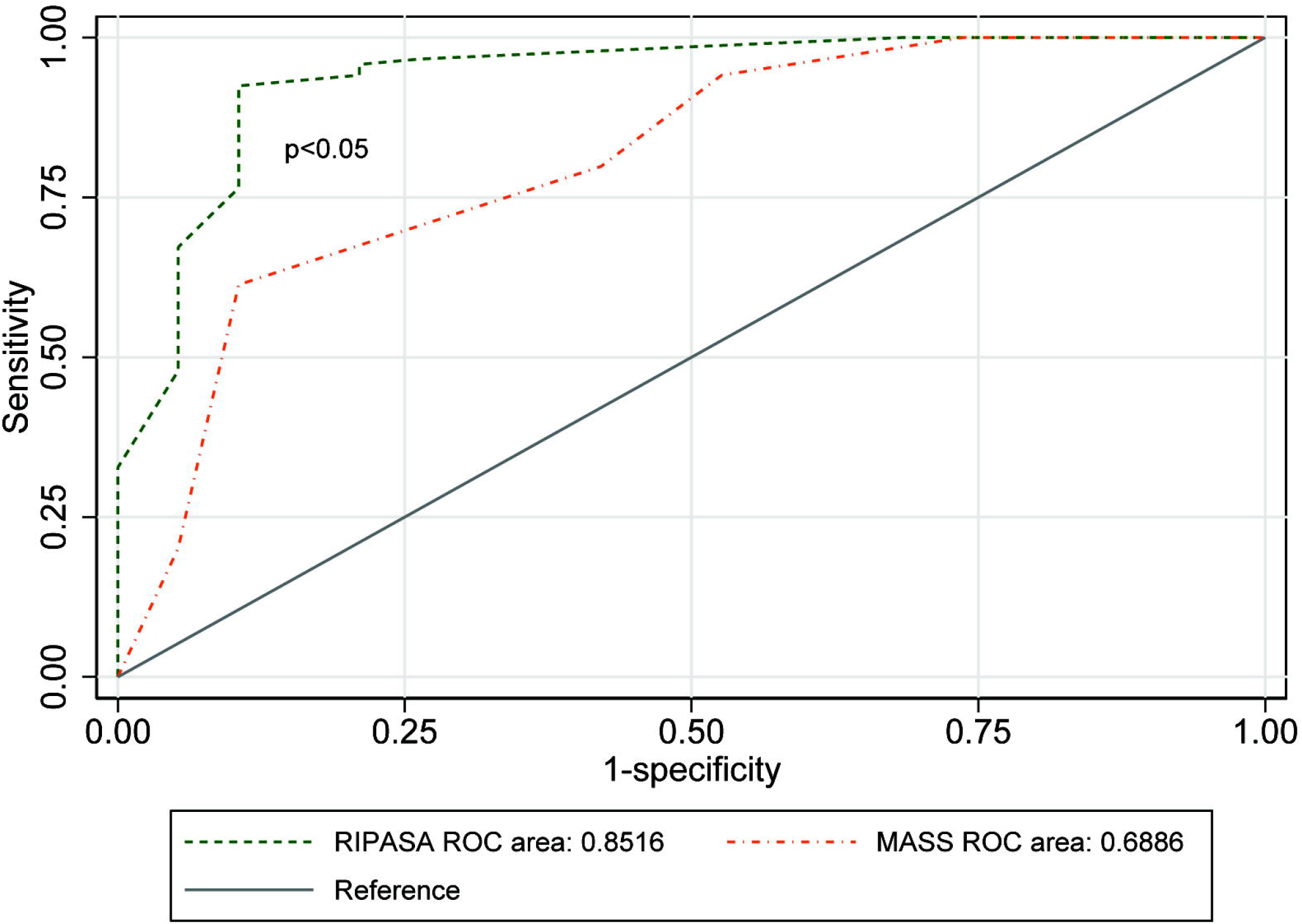
ROC-AUC plots for the MASS and RIPASA score model.

## Discussion

This study compared the diagnostic performance of the Modified Alvarado Scoring System (MASS) and the RIPASA scoring system for acute appendicitis in a peripheral district medical college hospital in Rajshahi, Bangladesh. Our analysis provides insights into their utility and limitations, offering a valuable perspective on their application in a specific demographic.

The demographic and clinical profile of our patient cohort, with a mean age of 26.2 years, is reflective of the global epidemiology of acute appendicitis, which predominantly affects individuals in this age group,[17] underscoring the relevance of our findings. The male predominance (55.0%) in our study corroborates the literature indicating a higher incidence of acute appendicitis among males.[18] Symptoms such as anorexia, nausea/vomiting, and the classic presentation of right iliac fossa (RIF) pain, alongside Rovsing’s sign, muscle guarding, and rebound tenderness, align with the established clinical manifestation of acute appendicitis.[19]

In evaluating the diagnostic scores against histopathological diagnosis as the gold standard, both the MASS and RIPASA scoring systems demonstrated notable sensitivity and specificity in distinguishing acute appendicitis from other conditions. The MASS system exhibited a sensitivity of 79.8%, correctly identifying a significant proportion of true positive cases, and a specificity of 57.9% for accurately discerning non-appendicitis cases. Comparing our study with previous research in Asian populations, the MASS model demonstrated a higher sensitivity of 79.8% compared to Al-Hashemy et al. (53.8%),[15] Jang et al. (50.6%),[20] and Chong et al. (68.3%)[9] but was close to Naeem et al. (83.3%),[16] and Shuaib et al. (82.8%).[21]. However, its specificity in our study (57.9%) was lower than reported in Al-Hashemy et al. (80%),[15] Jang et al. (94.5%),[20] and Chong et al. (87.9%),[9] but higher than Naeem et al. (41%),[16] and similar as Shuaib et al. (56%).[21] Conversely, the RIPASA score with a sensitivity of 96.6% and specificity of 73.7% performed consistently high across other studies. Our study reported a sensitivity of 96.6%, aligning closely with Singla et al. (95.6%),[8] Chong et al. (98%)[9] and Shuaib et al. (94.5%).[21] However, RIPASA’s specificity in our study (73.7%) was slightly lower than reported in Singla et al. (80%),[8] Chong et al. (81.3%),[9] and Shuaib et al. (88%).[21]

The higher sensitivity (96.6%) of the RIPASA score compared to the MASS system (79.8%) highlights its enhanced ability to correctly identify individuals with acute appendicitis. The specificity of the RIPASA score (73.7%), although higher than that of MASS (57.9%), suggests there is potential for further refinement in distinguishing non-appendicitis cases. The diagnostic advantage of the RIPASA scoring system in our study carries important clinical implications. Its elevated sensitivity indicates effectiveness in identifying acute appendicitis, potentially reducing the incidence of unnecessary surgeries in false-negative scenarios. The commendable specificity and positive predictive value also underscore its clinical utility. Moreover, both scoring systems showed reasonably high positive predictive values (PPV), with RIPASA marginally outperforming MASS. The notably higher negative predictive value (NPV) of RIPASA (77.8%) compared to MASS (31.4%) suggests that RIPASA is more dependable in ruling out appendicitis when the test result is negative. These outcomes collectively highlight the enhanced diagnostic accuracy of the RIPASA scoring system (93.5%) over the diagnostic accuracy of MASS (76.8%).

The Receiver Operating Characteristic (ROC) AUC analysis highlights the diagnostic advantage of the RIPASA scoring system over the Modified Alvarado Scoring System (MASS). Specifically, RIPASA’s ROC AUC is 23.7% higher than that of MASS, reflecting a statistically significant improvement in discriminatory capability (p < 0.05). This finding aligns with previous studies showing that the RIPASA scoring system provides superior diagnostic accuracy compared to MASS.[13], [14], [21], [22], [23]

This study has several strengths. It directly compares two widely used appendicitis scoring systems, the Modified Alvarado and RIPASA scores, providing valuable insights in a resource-limited setting. To our knowledge, this is the first study in Bangladesh to conduct such a comparison, utilizing histopathology as the diagnostic benchmark to ensure reliable confirmation of appendicitis cases. Additionally, the study’s focus on a low-resource environment enhances its relevance to similar settings. However, the study has some limitations. While twenty percent of the initially eligible patients were excluded from the final analysis, these exclusions were necessary to maintain diagnostic consistency and accuracy, which could limit the generalizability of the findings to cases without histopathology or those treated conservatively. The study was conducted in a single hospital, which also limits its generalizability. Variability in clinical assessments by different attending surgeons could result in observer bias.

## Conclusion

This study provides valuable insights into the diagnostic performance of the Modified Alvarado and RIPASA scoring systems for acute appendicitis in a specific population. The RIPASA scoring system exhibited superior diagnostic performance across several metrics, including sensitivity, specificity, positive and negative predictive values, overall diagnostic accuracy, and the AUC, when compared to the MASS system. These results enrich the ongoing discourse on diagnostic strategies for acute appendicitis, highlighting the RIPASA scoring system’s value in similar healthcare settings where access to advanced diagnostic resources may be limited.

Future research endeavors should focus on validating these findings in diverse populations and settings. Additionally, prospective studies could explore the integration of advanced imaging modalities and biomarkers to enhance diagnostic accuracy further and can develop a new scoring model which is more appropriate for the local context. Comparative analyses with other scoring systems and clinical decision support tools would contribute to a more comprehensive understanding of the optimal approach to acute appendicitis diagnosis.

## Acknowledgments

This paper utilized data from Master’s thesis of Nafisa conducted by the Rajshahi Medical College & Hospital, Rajshahi, Bangladesh under the Bangabandhu Sheikh Mujib Medical University (BSMMU), Bangladesh. We extend our sincere gratitude to the participants and the faculty members of the Department of General Surgery, Pathology, and Biochemistry at Rajshahi Medical College & Hospital, Rajshahi, Bangladesh, for their invaluable contributions to this study.

## Author Contributions

Nafisa, Shahid, Baharul, and Monower conceptualized and designed the study. Nafisa led the study implementation, with data collection conducted by Nafisa, Momena, and Tanvir. Nafisa prepared the initial manuscript draft, while Monower performed the statistical analyses and interpreted the results. Nafisa and Monower jointly contributed to the interpretation of findings and the literature review. Baharul supervised the thesis work.

## Data availability statement

The data supporting the findings of this study are openly available in Zenodo at https://doi.org/10.5281/zenodo.14043746, under the citation: Naz, N. (2024). A Comparative Study of RIPASA and Modified Alvarado Scoring System for the Diagnosis of Acute Appendicitis [Data set]. Access to the dataset requires prior permission from the corresponding author for reuse.

## Supporting information

Supplementary figure 1: Flow diagram of participants

Supplementary tables 1: Modified Alvarado Scoring System (MASS)

Supplementary tables 2: Score distribution of RIPASA score system

## References

[1] M. F. Ditillo, J. D. Dziura, and R. Rabinovici, “Is it safe to delay appendectomy in adults with acute appendicitis?,” Ann Surg, vol. 244, no. 5, pp. 656–660, Nov. 2006, doi: 10.1097/01.sla.0000231726.53487.dd.

[2] Chong C F, MW W Adi, Thien A, and Suyoi A, “Development of the RIPASA score: a new appendicitis scoring system for the diagnosis of acute appendicitis.,” Singapore Med J, vol. 51, no. 3, pp. 220–225, 2010.

[3] Delibegovic Samir, “Acute Appendicitis: Diagnosis and Treatment, with Special Attention to a Laparoscopic Approach,” Emerg Med (Los Angel), vol. 05, no. 03, 2015, doi: 10.4172/2165-7548.1000255.

[4] R. R. Gorter et al., “Diagnosis and management of acute appendicitis. EAES consensus development conference 2015,” Surg Endosc, vol. 30, no. 11, pp. 4668–4690, Nov. 2016, doi: 10.1007/s00464-016-5245-7.

[5] M. Korkut, C. Bedel, Y. Karanci, A. Avci, and M. Duyan, “Accuracy of Alvarado, Eskelinen, Ohmann, RIPASA and Tzanakis Scores in Diagnosis of Acute Appendicitis; a Cross-sectional Study,” Archives of Academic EmergencyMedicine, vol. 8, no. 1, p. e20, Mar. 2020.

[6] Alvarado A, “A Practical Score for the Early Diagnosis of Acute Appendicitis,” Ann Emerg Med, vol. 15, no. 5, pp. 557–564, 1986, doi: 10.1016/s0196-0644(86)80993-3.

[7] E. S. Kanumba, J. B. Mabula, P. Rambau, and P. L. Chalya, “Modified Alvarado Scoring System as a diagnostic tool for Acute Appendicitis at Bugando Medical Centre, Mwanza, Tanzania,” BMC Surg, vol. 11, 2011, doi: 10.1186/1471-2482-11-4.

[8] A. Singla, S. Singla, M. Singh, and D. Singla, “A comparison between modified Alvarado score and RIPASA score in the diagnosis of acute appendicitis,” Updates Surg, vol. 68, no. 4, pp. 351–355, Dec. 2016, doi: 10.1007/s13304-016-0381-0.

[9] C. F. Chong et al., “Comparison of RIPASA and Alvarado scores for the diagnosis of acute appendicitis.,” Singapore Med J, vol. 52, no. 5, pp. 340–5, May 2011.

[10] H. Erdem et al., “Alvarado, Eskelinen, Ohhmann and Raja Isteri Pengiran Anak Saleha appendicitis scores for diagnosis of acute appendicitis,” World J Gastroenterol, vol. 19, no. 47, pp. 9057–9062, Dec. 2013, doi: 10.3748/wjg.v19.i47.9057.

[11] Butt MQ, Chatha SS, Ghumman AQ, and Farooq M, “RIPASA Score: A New Diagnostic Score for Diagnosis of Acute Appendicitis,” J Coll Physicians Surg Pak, vol. 24, no. 12, pp. 894–897, 2014.

[12] Nafisa Naz, “A Comparative Study of RIPASA and Modified Alvarado Scoring System for the Diagnosis of Acute Appendicitis [Data set],” Zenodo, Nov. 2024, doi: 10.5281/zenodo.14043746.

[13] N. Damburacı, B. Sevinç, M. Güner, and Ö. Karahan, “Comparison of Raja Isteri Pengiran Anak Saleha Appendicitis and modified Alvarado scoring systems in the diagnosis of acute appendicitis,” ANZ J Surg, vol. 90, no. 4, pp. 521–524, Apr. 2020, doi: 10.1111/ans.15607.

[14] S. Noor, A. Wahab, G. Afridi, and K. Ullah, “Comparing Ripasa Score And Alvarado Score In An Accurate Diagnosis Of Acute Appendicitis.,” J Ayub Med Coll Abbottabad, vol. 32, no. 1, pp. 38– 41, 2020.

[15] A. M. Al-Hashemy and M. I. Seleem, “Appraisal of the modified Alvarado Score for acute appendicits in adults.,” Saudi Med J, vol. 25, no. 9, pp. 1229–31, Sep. 2004.

[16] M. T. Naeem et al., “Diagnostic accuracy of Alvarado scoring system relative to histopathological diagnosis for acute appendicitis: A retrospective cohort study,” Annals of Medicine and Surgery, vol. 81, Sep. 2022, doi: 10.1016/j.amsu.2022.104561.

[17] Y. J. Noudeh, N. Sadigh, and A. Y. Ahmadnia, “Epidemiologic features, seasonal variations and false positive rate of acute appendicitis in Shahr-e-Rey, Tehran,” International Journal of Surgery, vol. 5, no. 2, pp. 95–98, Apr. 2007, doi: 10.1016/j.ijsu.2006.03.009.

[18] G. Y. Stein et al., “Sex differences in the epidemiology, seasonal variation, and trends in the management of patients with acute appendicitis,” Langenbecks Arch Surg, vol. 397, no. 7, pp. 1087–1092, Oct. 2012, doi: 10.1007/s00423-012-0958-0.

[19] F. Dixon and A. Singh, “Acute appendicitis,” Surgery (United Kingdom), vol. 38, no. 6, pp. 310– 317, Jun. 2020, doi: 10.1016/j.mpsur.2020.03.015.

[20] S. O. Jang, B. S. Kim, and D. J. Moon, “Application of alvarado score in patients with suspected appendicitis,” Korean J Gastroenterol, vol. 52, no. 1, pp. 27–31, Jul. 2008.

[21] A. Shuaib, A. Shuaib, Z. Fakhra, B. Marafi, K. Alsharaf, and A. Behbehani, “Evaluation of modified Alvarado scoring system and RIPASA scoring system as diagnostic tools of acute appendicitis,” World J Emerg Med, vol. 8, no. 4, p. 276, 2017, doi: 10.5847/wjem.j.1920-8642.2017.04.005.

[22] N. Heiranizadeh, S. M. H. Mousavi Beyuki, S. kargar, A. Abadiyan, and H. R. Mohammadi, “Alvarado or RIPASA? Which one do you use to diagnose acute appendicitis?: A cross-sectional study,” Health Sci Rep, vol. 6, no. 1, Jan. 2023, doi: 10.1002/hsr2.1078.

[23] M. M. Chisthi, A. Surendran, and J. T. Narayanan, “RIPASA and air scoring systems are superior to alvarado scoring in acute appendicitis: Diagnostic accuracy study,” Annals of Medicine and Surgery, vol. 59, pp. 138–142, Nov. 2020, doi: 10.1016/j.amsu.2020.09.029.

