## Supporting information for "Diagnosing Appendicitis with Precision: A Comparative Analysis of Modified Alvarado and RIPASA Scoring Systems in a Northern Divisional Hospital of Bangladesh"

**Supplementary figure**

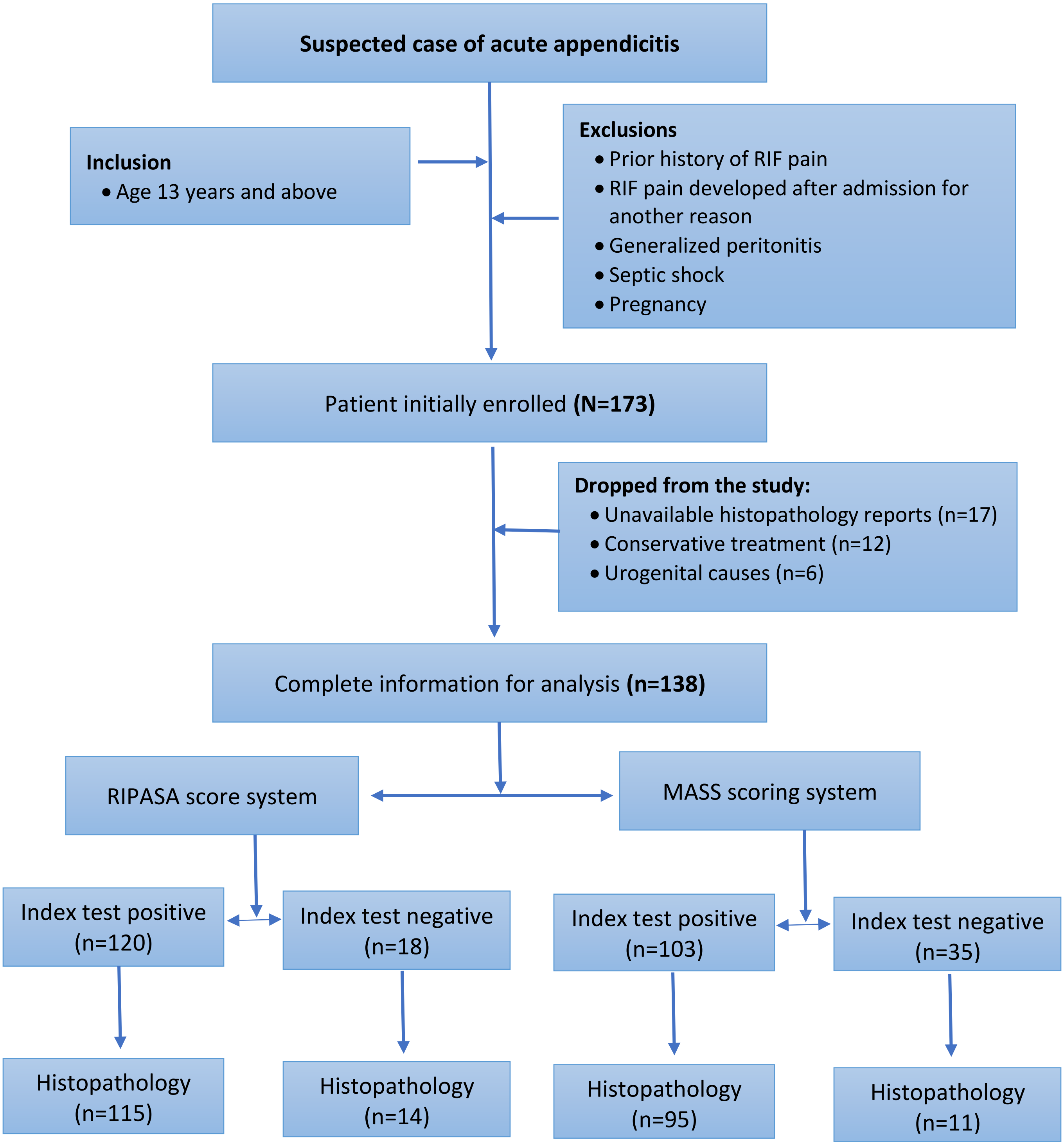


Supplementary figure 1: Flow diagram of participants

**Supplementary tables**

Supplementary tables 1: Modified Alvarado Scoring System (MASS)

| | **FEATURE** | **SCORE** |
|---|---|---|
| | Migratory RIF pain | 1 |
| Symptoms | Anorexia | 1 |
| | Nausea/ Vomiting | 1 |
| | Tenderness in RIF | 2 |
| | Rebound tenderness in RIF | 1 |
| Signs | Elevated temperature | 1 |
| Laboratory | Leukocytosis | 2 |
| | **TOTAL** | **9** |

Supplementary tables 2: Score distribution of RIPASA score system

| PATIENT'S DEMOGRAPHIC | SCORE |
|---|---|
| Female | 0.5 |
| Male | 1.0 |
| Age< 39.9years | 1.0 |
| Age> 40years | 0.5 |
| **SYMPTOMS** | |
| RIF pain | 0.5 |
| Pain migration to RIF | 0.5 |
| Anorexia | 1.0 |
| Nausea & vomiting | 1.0 |
| Duration of symptoms <48hrs | 1.0 |
| Duration of symptoms >48hrs | 0.5 |
| **SIGNS** | |
| RIF tenderness | 1.0 |
| Guarding | 2.0 |
| Rebound tenderness | 1.0 |
| Rovsing's sign | 2.0 |
| Fever>$37^0$C, <$39^0$C | 1.0 |
| **INVESTIGATIONS** | |
| Raised WBC count | 1.0 |
| Negative urinalysis | 1.0 |
| **ADDITIONAL SCORES** | |
| Foreign NRIC | 1.0 |
